# Image-based Mandibular and Maxillary Parcellation and Annotation using Computer Tomography (IMPACT): A Deep Learning-based Clinical Tool for Orodental Dose Estimation and Osteoradionecrosis Assessment

**DOI:** 10.1101/2025.03.18.25324199

**Authors:** Laia Humbert-Vidan, Austin H. Castelo, Renjie He, Lisanne V. van Dijk, Dong Joo Rhee, Congjun Wang, He C. Wang, Kareem A Wahid, Sonali Joshi, Parshan Gerafian, Natalie West, Zaphanlene Kaffey, Sarah Mirbahaeddin, Jaqueline Curiel, Samrina Acharya, Amal Shekha, Praise Oderinde, Alaa M.S. Ali, Andrew Hope, Erin Watson, Ruth Wesson-Aponte, Steven J. Frank, Carly E.A. Barbon, Kristy K. Brock, Mark S. Chambers, Muhammad Walji, Katherine A. Hutcheson, Stephen Y. Lai, Clifton D. Fuller, Mohamed A. Naser, Amy C. Moreno, the OPC-SURVIVOR Program, the MD Anderson Head and Neck Cancer Symptom Working Group

**Author notes:** co-corresponding authors **Corresponding author:** Amy C. Moreno, MS MD, Assistant Professor, H&N Service, Department of Radiation Oncology, UT MD Anderson Cancer Center.

## Abstract

**Background:** Accurate delineation of orodental structures on radiotherapy CT images is essential for dosimetric assessments and dental decisions. We propose a deep-learning auto-segmentation framework for individual teeth and mandible/maxilla sub-volumes aligned with the ClinRad ORN staging system.

**Methods:** Mandible and maxilla sub-volumes were manually defined, differentiating between alveolar and basal regions, and teeth were labelled individually. For each task, a DL segmentation model was independently trained. A Swin UNETR-based model was used for the mandible sub-volumes. For the smaller structures (e.g., teeth and maxilla sub-volumes) a two-stage segmentation model first used the ResUNet to segment the entire teeth and maxilla regions as a single ROI that was then used to crop the image input of the Swin UNETR. In addition to segmentation accuracy and geometric precision, a dosimetric comparison was made between manual and model-predicted segmentations.

**Results:** Segmentation performance varied across sub-volumes – mean Dice values of 0.85 (mandible basal), 0.82 (mandible alveolar), 0.78 (maxilla alveolar), 0.80 (upper central teeth), 0.69 (upper premolars), 0.76 (upper molars), 0.76 (lower central teeth), 0.70 (lower premolars), 0.71 (lower molars) – and exhibited limited applicability in segmenting teeth and sub-volumes often absent in the data. Only the maxilla alveolar central sub-volume showed a statistically significant dosimetric difference (Bonferroni-adjusted p-value = 0.02).

**Conclusion:** We present a novel DL-based auto-segmentation framework of orodental structures, enabling spatial localization of dose-related differences in the jaw. This tool enhances image-based bone injury detection, including ORN, and improves clinical decision-making in radiation oncology and dental care for head and neck cancer patients.

## 1. Introduction

The introduction of modern imaging and radiation therapy techniques has enabled highly conformal irradiation of head and neck tumors but, due to the close proximity of critical structures in that region, non-target organs such as the mandible or maxilla inevitably absorb ionizing radiation, resulting in detrimental complications such as osteoradionecrosis (ORN). Radiation-induced injury to the mandible and maxilla results from direct cellular and vascular damage to the bone, resulting in reduced blood supply and subsequent necrosis of the bone tissue (i.e., ORN). Thus, the healing process occurring after a dental extraction or any physical trauma to the bone may be slower or absent in irradiated bone^1^. Such considerations are important in dental (e.g., dental extractions or implants) and oncological (e.g., radiation dose distribution) decisions made pre-, during, and post-treatment of patients with head and neck cancer^2–4^.

Accurate delineation of orodental structures on radiotherapy computed tomography (CT) images is essential for effective dosimetric evaluation during treatment planning optimization and also post-treatment to support dental decisions, such as evaluating the feasibility of dental implants or assessing the risk of normal tissue damage following irradiation. Manual segmentation of orodental structures is a time-consuming, poorly reproducible and often challenging task due to strong artifacts caused by high density (higher attenuation) materials such as dental fillings, restorations or titanium implants, which result in blurred edges of the structure^5^. Boundaries between adjacent teeth are often indistinct, the angles of teeth may diverge from crown to root, and positional variations, such as gaps or missing teeth, can further complicate the process. This has led to increased research focus in recent years of developing automated segmentation methods for orodental structures, mostly mandible^5–7^ and teeth^8–13^.

These studies, however, considered the mandible as a single structure disregarding its heterogeneity with regards to composition and radiobiological characteristics. The bone composition varies across the mandible and maxilla, with density, metabolic and radiosensitivity differences between sub-volumes. For instance, the alveolar process surrounding the teeth has a higher proportion of spongy or trabecular bone, which is more vascularized and metabolically active and is more sensitive to radiation as it contains bone marrow^14^. The basal region has a denser composition, with a larger proportion of cortical bone^15^.

On the other hand, most teeth auto-segmentation studies leverage cone-beam CT data, which is considered the gold standard for volumetric dental imaging. Limited series have focused on teeth auto-segmentation specifically to support radiation oncology for head and neck cancer patients utilizing surveillance contrast-enhanced CT images and/or radiation simulation CT, which pose additional challenges due to lower spatial resolution in bone compared to cone-beam CT yet are more clinically relevant for this patient population.

In this study, we present a novel and comprehensive approach to the auto-segmentation of bony odontic structures on CT images, aimed at supporting clinical decision-making in radiation therapy for patients with head and neck cancer. Advancing beyond existing whole mandible auto-segmentation methods^5–8^, we propose a refined definition of mandible and maxilla sub-volumes that accounts for variations in bone composition and the anatomical and physiological differences within these structures. Our proposed sub-volumes are designed to align with the recently ASCO-endorsed ORN staging system, the ClinRad system^16^, which incorporates radiological assessment of the vertical extent of bone damage. This alignment has the potential to improve early detection of mandible and maxilla damage, particularly in cases with intact mucosa, providing a more clinically meaningful segmentation framework.

## 2. Materials and Methods

### 2.1. Patients

After institutional review board approval (RCR030800), data for 60 cases from a philanthropically funded observational cohort (Stiefel Oropharynx Cancer Cohort, PA14-0947) were extracted retrospectively. Cases with more than 20 missing teeth or with overt image artifacts (i.e., streak artifact preventing from accurate manual teeth delineation) were excluded. Radiotherapy treatment data included planning CT images, structure sets and planned radiation dose distributions. Clinical treatment plans had been created using the RayStation, Eclipse and Pinnacle treatment planning systems.

### 2.2. Contouring

#### Mandible and maxilla sub-volumes

Mandible and maxilla sub-volumes were manually contoured on planning CT images in RayStation treatment planning system (RaySearch Laboratories, Sweden) to include left/right/central alveolar and basal bone regions, i.e., a total of 12 sub-volumes were obtained per case (Figure 1). Laterality of the sub-volumes was defined by grouping the teeth into three sextants. A detailed description of the sub-volume contouring process is provided in Supplement A. The alveolar region was defined as an expansion from the alveolar crest of 5 mm inferiorly and 3 mm superiorly for the mandible and maxilla, respectively^15^. To ensure that our model aligns with the ClinRad ORN classification system, i.e., to allow for a differentiation between alveolar and basal regions, the contours of the lateral alveolar sub-volumes at the molars level of the mandible were manually adjusted to meet a ‘contracted’ mandible contour that was defined by reducing the mandible bone contour by 4 mm. This allowed the basal region to continuously expand from the chin to the condyles through the angle (see supplementary Figure A1).

**Figure 1.**
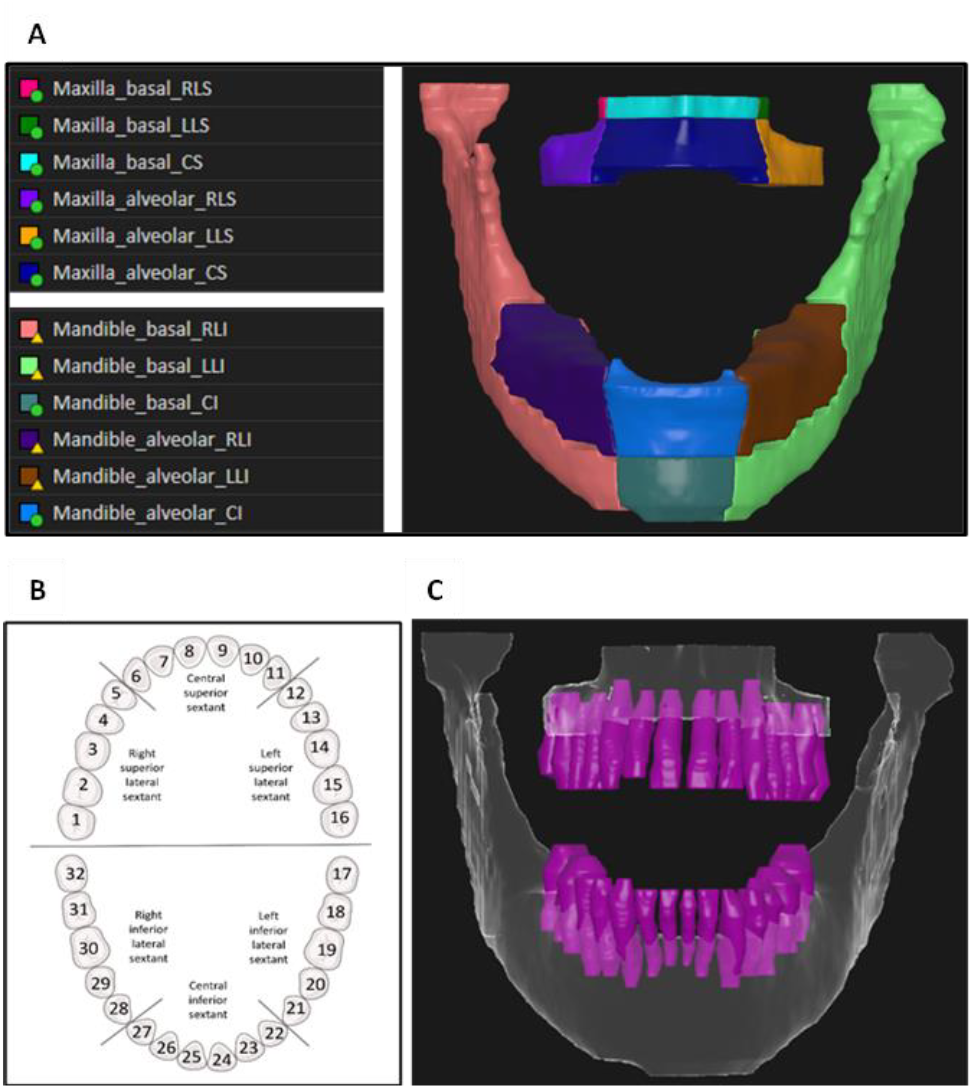
Illustration of the orodental structures definitions utilized: A) maxilla and mandible sub-volumes defining left/right/central alveolar and basal bone regions, where laterality of the sub-volumes was defined by grouping the teeth into three sextants (B), and C) individual teeth segmentation including the roots extending into the mandible and maxilla bones, where teeth were numbered following the Universal Numbering System (1-32).

#### Teeth

Individual teeth were manually contoured and numbered following the American Dental Association (ADA) Universal Numbering System (1-32) (Figure 1). Missing teeth were not contoured but accounted for with regards to the numbering; a tooth was considered as missing if the gap between adjacent teeth was larger than 5 mm. To ensure generalizability of the model on edentulous or semi-edentulous patients, empty tooth sockets in cases with missing teeth were included in the alveolar region by manually adjusting the teeth expansion contour (e.g., Supplementary Figure A2).

### 2.3. Image preprocessing

DICOM (Digital Imaging and Communications in Medicine) files (CT images and RT structure set) were exported from the radiotherapy treatment planning system and converted into NIfTI (Neuroimaging Informatics Technology Initiative) format using *SimpleITK*^17^ and *dcmrtstruct2nii* software^18^. Structures were converted into masks and assigned individual labels; a priority-based approach (with central volumes having priority over lateral ones) was used for mandible and maxilla sub-volumes to address potential voxel overlap during the manual contouring process. All images were resampled to a fixed resolution of 1 mm in the axial plane (x, y dimensions) and 2 mm in the z dimension, with CT Hounsfield Units (HU) clipped between −200 and 1600 to enhance bone structures and rescaled between 0 and 1.

### 2.4. Model implementation

The anatomy of the different orodental structures considered varies significantly (e.g., individual teeth are significantly smaller than mandible or maxilla sub-volumes and the maxilla basal region is also smaller to that of the mandible), which led to the decision of addressing the following three segmentation tasks separately: mandible sub-volumes, maxilla sub-volumes and individual teeth. For each task, a deep learning (DL) segmentation model was independently trained. We used the Residual U-Net (ResUNet)^19^ and Swin UNETR^20^ Transformer-based convolutional neural networks from the Medical Open Network for

Artificial Intelligence (MONAI) software package^21^. A one-stage Swin UNETR-based model was used for the mandible sub-volumes. For the smaller structures, such as teeth and maxilla sub-volumes, a two-stage segmentation model was developed to enhance segmentation performance in the presence of class imbalance^22^. In the two-stage model, ResUNet was used in the first stage to segment the entire teeth and maxilla regions as a single ROI. The center of mass of this ROI was then used to crop the image into a fixed voxel size of 128×128×128, which was subsequently input into the Swin UNETR model in the second stage for precise segmentation of individual teeth and maxilla sub-volumes.

Data augmentation included random rotations (±12°), scaling (±10%), and random intensity scaling and shifting (±10%). Both the one-stage and two-stage models were trained for 500 epochs with a learning rate of 10^−4^ using the Dice-Cross Entropy loss function from MONAI. A 5-fold cross-validation approach was employed, with 50 cases for training and 10 for testing. For testing, five sub-models for each task (mandible, maxilla, and teeth) were applied, and the final segmentation was obtained using a majority vote across the outputs of these models.

### 2.5. Model performance

Patients were randomly split into train (n=50) and test (n=10) subsets. Model performance was evaluated on the test subset with regards to segmentation accuracy and geometric precision using the Dice Similarity Coefficient (Dice), Mean Intersection over Union (mIoU), 95^th^ percentile Hausdorff Distance (HD95) and Average Surface Distance (ASD) metrics. Dice and mIoU provide a measure of overlap between predicted segmentations and ground truth, ranging from 0 (no overlap) to 1 (perfect overlap). HD95 and ASD capture segmentation boundary precision, with smaller distances corresponding to better alignment between the predicted and ground truth surfaces, with HD95 specifically capturing the 95th percentile of surface distances to reduce sensitivity to outliers. These metrics were calculated in Python using the packages *medpy*.*metric*.*binary, scipy*.*spatial*.*distance, surface_distance*.*metrics* and *NumPy*.

### 2.6. Dosimetric comparison

The performance of the segmentation models was further assessed by comparing the mean (Dmean) and maximum (Dmax) dose values within each segmented class for both manual (i.e., ground truth) and model-predicted segmentations. The analysis involved several steps: loading NIfTI images of the radiation dose and segmentations, resampling images to ensure alignment, masking the radiation dose distribution image, and calculating dose metrics for the distinct anatomical sub-regions segmented. The Wilcoxon signed-rank test (with Bonferroni correction for multiple comparisons) was used to compare Dmax and Dmean values across all classes to assess statistically significant differences.

## 3. Results

### 3.1. Patients

The final cohort consisted of 60 patients treated with head and neck cancer who underwent radiotherapy with prescribed doses from 60 Gy in 30 fractions to 70 Gy in 35 fractions. Table 1 summarizes the demographic and dental characteristics of this cohort, specifically for the train and test subsets.

**Table 1.**
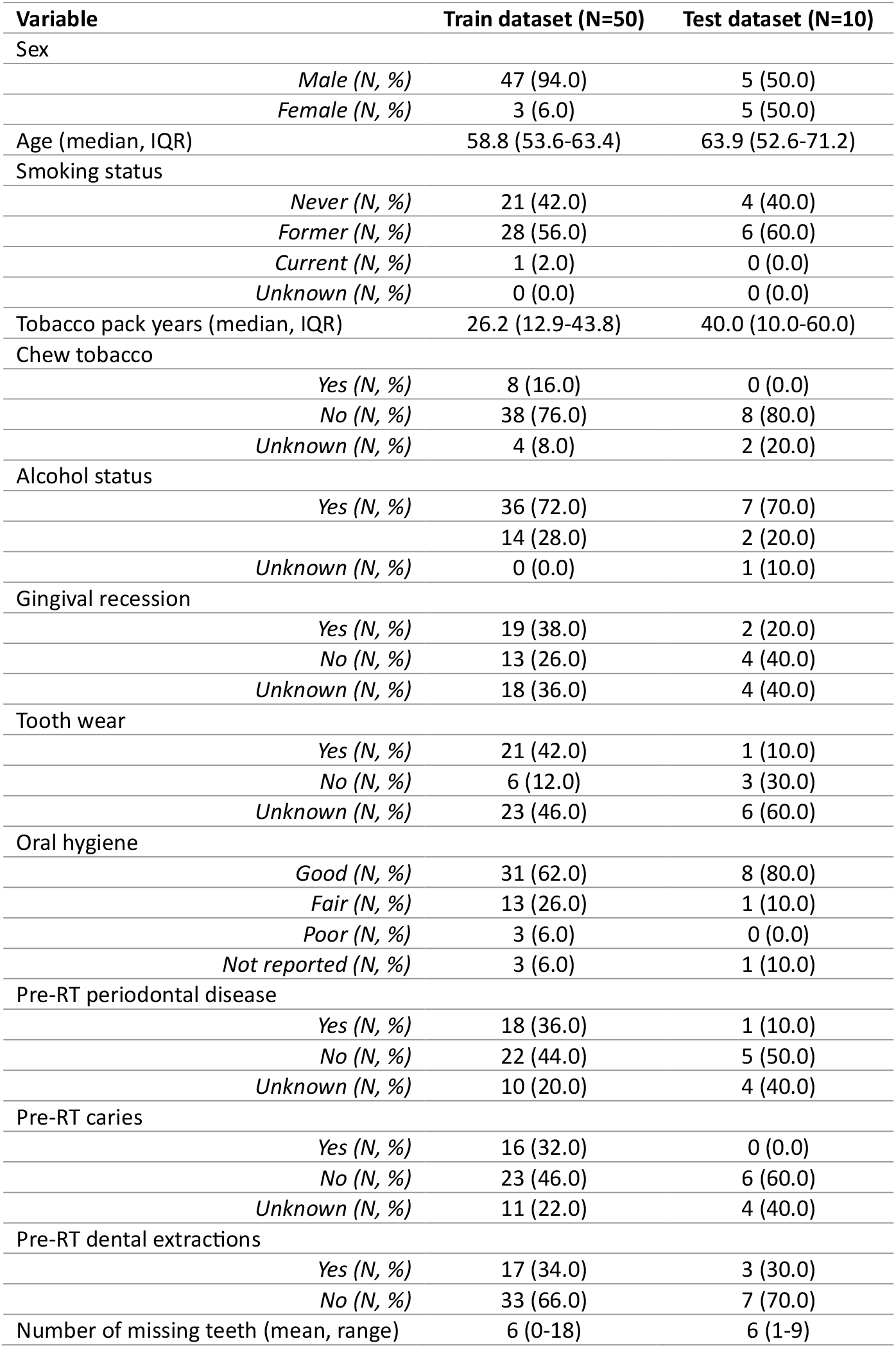
Cohort demographic and dental characteristics.

### 3.2. Model performance

Successful segmentation of mandible sub-volumes was achieved, with basal sub-volumes showing overall higher segmentation accuracy compared to alveolar sub-volumes (Figure 2a). In comparison to the mandible, the segmentation performance for maxilla sub-volumes showed notable differences (Figure 2b). While the mandible basal sub-volumes achieved high accuracy, maxilla basal sub-volumes were not successfully segmented, most likely due to limited training data for this often small and even non-existing sub-volume. However, both maxilla and mandible alveolar sub-volumes exhibited low average surface distances, indicating consistent precision in boundary delineation. Full quantitative metrics are provided in Table 2.

**Table 2.** Mandible and maxilla sub-volumes and teeth segmentation models’ segmentation performance results. Mandible and maxilla sub-volumes were grouped by the following regions: basal (classes 1-3) vs. alveolar (classes 4-6). Teeth were grouped as follows: Upper Central (6-11), Upper Premolars (4, 5, 12, 13), Upper Molars (1-3, 14-16), Lower Central (22-27), Lower Premolars (20, 21, 28, 29) and Lower Molars (17-19, 30-32).

| Anatomical region | Dice | mIoU | HD (mm) | ASD (mm) |
| --- | --- | --- | --- | --- |
|  | Mean (range) | Mean (range) | Mean (range) | Mean (range) |
| Mandible basal | 0.85 (0.80-0.88) | 0.74 (0.67-0.79) | 1.17 (1.00-2.03) | 0.30 (0.13-1.38) |
| Mandible alveolar | 0.82 (0.69-0.89) | 0.70 (0.53-0.80) | 1.52 (1.00-3.24) | 0.24 (0.12-0.52) |
| Maxilla alveolar | 0.78 (0.64-0.86) | 0.64 (0.47-0.75) | 1.56 (1.00-2.74) | 0.28 (0.15-0.54) |
| Upper central teeth | 0.80 (0.53-0.89) | 0.69 (0.39-0.81) | 1.43 (1.00-3.09) | 0.27 (0.11-0.92) |
| Upper premolars | 0.69 (0.37-0.87) | 0.56 (0.25-0.76) | 2.30 (1.00-5.14) | 0.62 (0.14-2.18) |
| Upper molars | 0.76 (0.40-0.90) | 0.64 (0.27-0.82) | 3.65 (1.00-16.49) | 0.57 (0.10-2.28) |
| Lower central teeth | 0.76 (0.42-0.89) | 0.63 (0.28-0.80) | 1.62 (1.00-3.73) | 0.34 (0.11-1.10) |
| Lower premolars | 0.71 (0.38-0.86) | 0.57 (0.25-0.76) | 2.32 (1.00-5.07) | 0.51 (0.14-1.60) |
| Lower molars | 0.63 (0.38-0.75) | 0.54 (0.28-0.68) | 3.61 (2.26-8.03) | 1.35 (0.63-2.78) |

**Figure 2.**
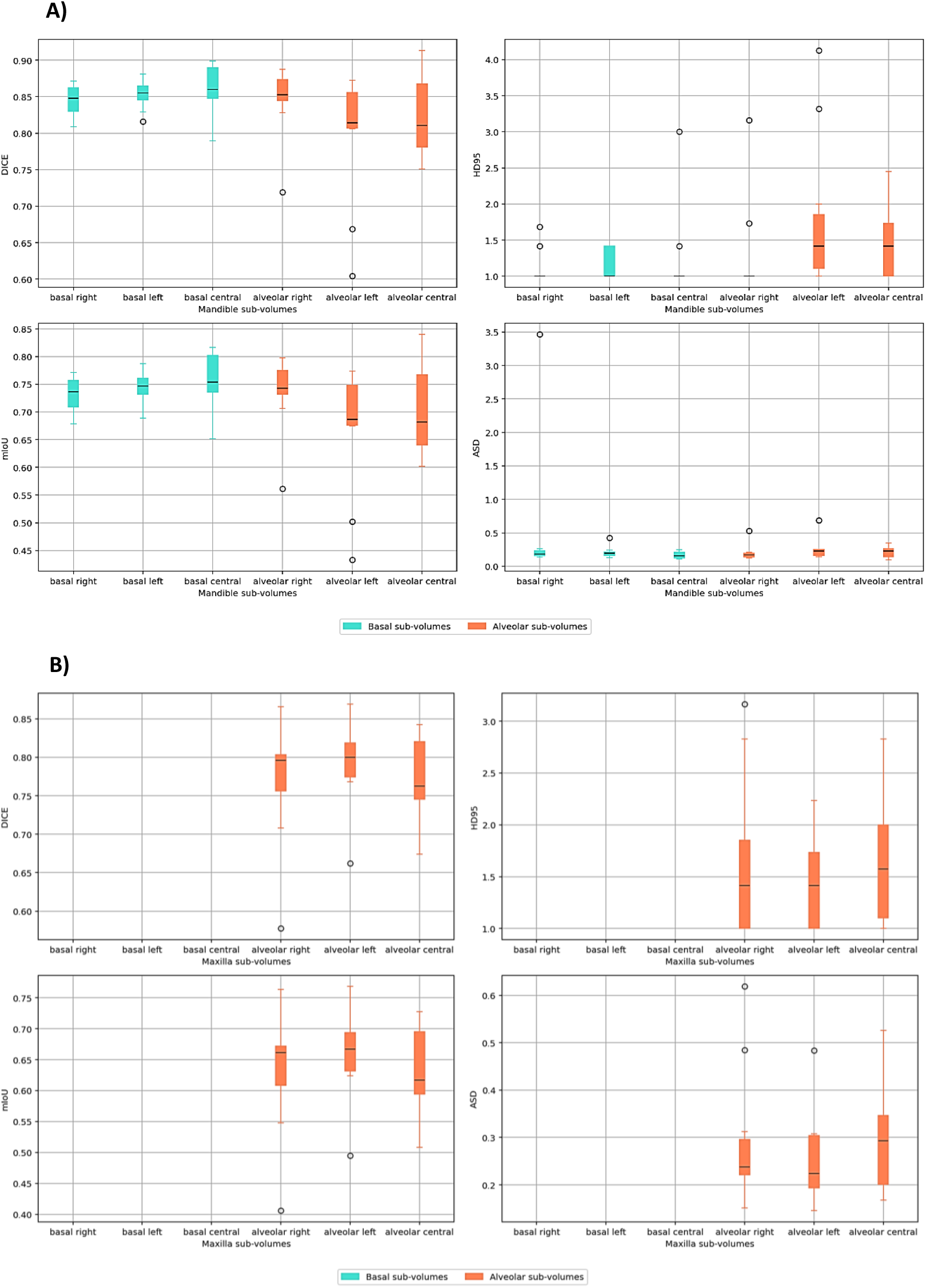
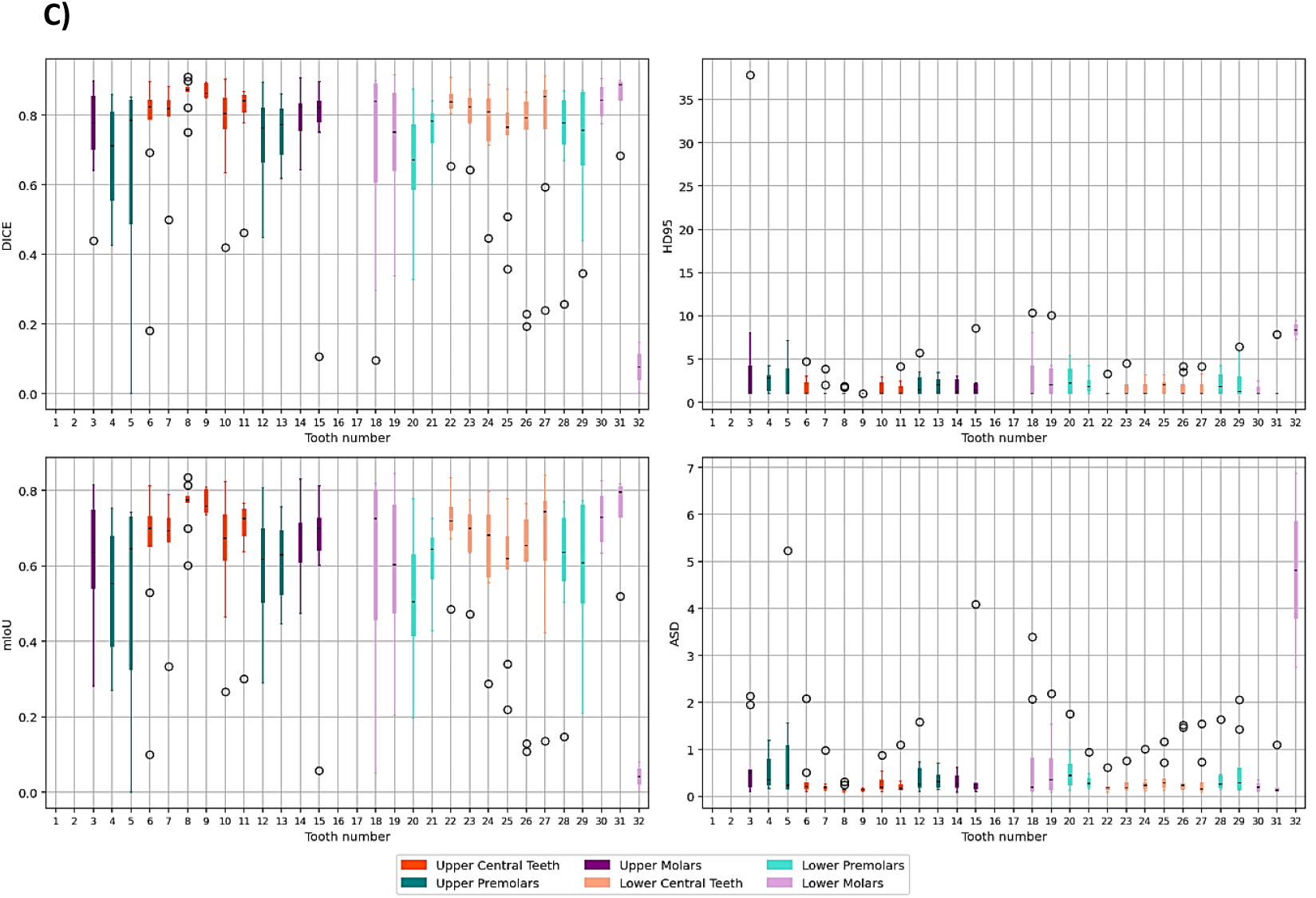
Boxplots describing the distribution of performance metrics by class for the A) mandible sub-volumes B) maxilla sub-volumes and C) teeth segmentation models.

The teeth segmentation model demonstrated variable performance across different tooth groups (Figure 2c), which can be partially attributed to the proportion of missing teeth in the datasets (see Supplement B). Central teeth, both upper and lower, which achieved higher segmentation accuracy, also had lower percentages of missing data in both the training and test datasets. In contrast, premolars and molars, particularly upper molars, exhibited higher percentages of missing data, correlating with their lower Dice scores and mIoU values. Central teeth, both upper and lower, achieved higher segmentation accuracy compared to premolars and molars (Figure 2c).

### 3.3. Dosimetric performance

Figure 3 shows a comparison between predicted class segmentations and manually contoured structures for 8 cases (two of the test cases had radiation dose fields far from the jaws with no dose delivered to the structures of interest). While most cases fell within the ±2.5 Gy dose difference range for Dmean in the mandible and maxilla sub-volumes, more outliers were observed for Dmax. The Dmean relative differences observed ranged from −9.4% to 3.4% and −7.1% to 6.5% for the mandible and the maxilla, respectively. For Dmax, these ranges were wider: −10.3% to 24.5% (mandible) and −34.0% to 8.4% (maxilla). For teeth, the large majority of the dosimetric outliers were observed for the molar and premolars. The Dmean relative differences observed ranged from −7.1% to 9.7%, −11.5% to 10.6%, and −12.6% to 4.5% for the central teeth, premolars and molars, respectively. For Dmax, these ranges were also wider: −7.8% to 30.0% (central teeth), −9.0% to 24.1% (premolars) and −6.1% to 82.4% (molars). However, after applying a Bonferroni correction for multiple comparisons to the Wilcoxon signed-rank test results, only the dosimetric difference observed for the maxilla alveolar central sub-volume was found to be statistically significant (adjusted p-value = 0.02).

**Figure 3.**
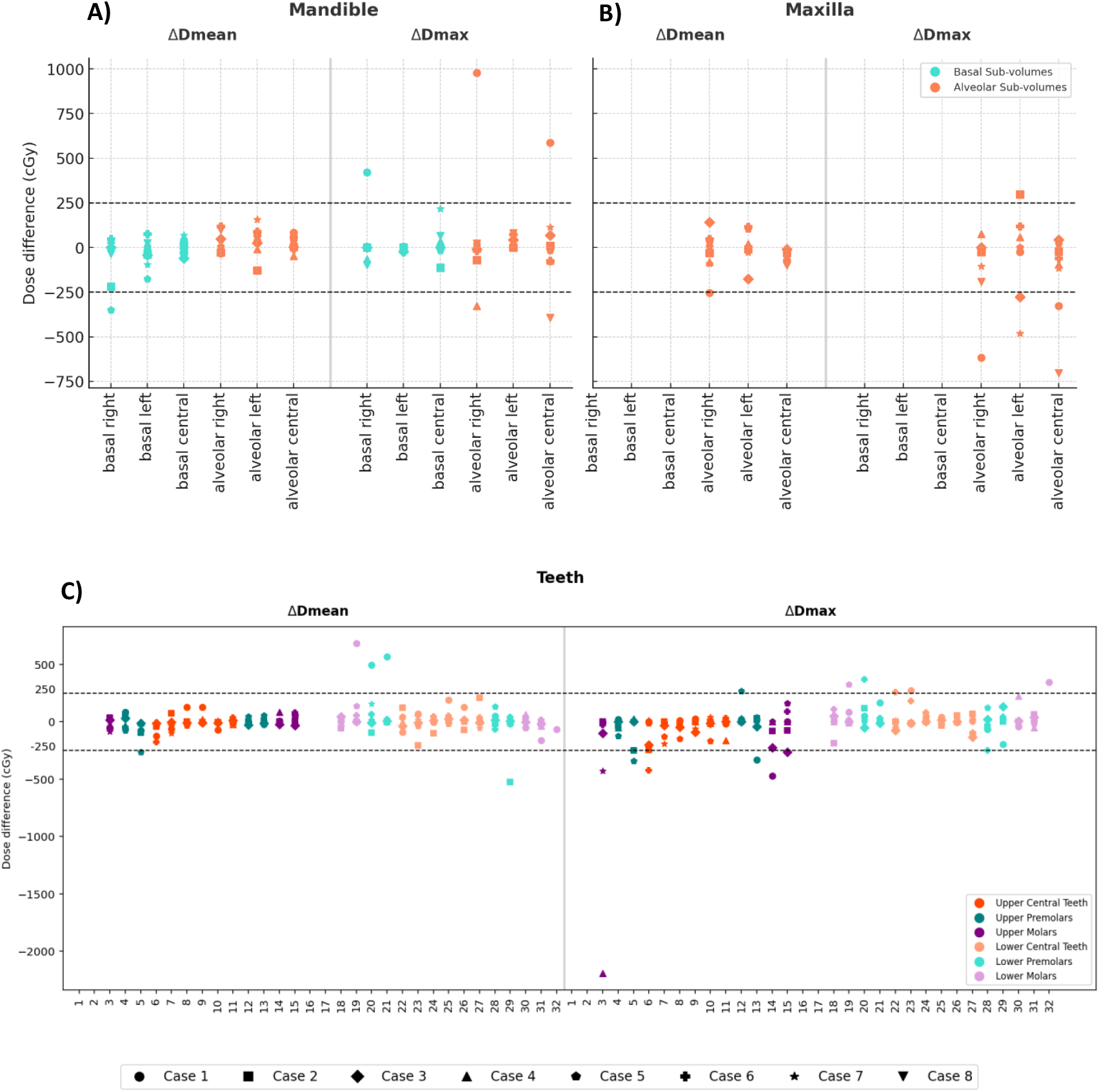
Differences in dosimetric performance (ΔDmax and ΔDmean) between predicted and ground truth orodental structures – mandible sub-volumes (A), maxilla sub-volumes (B) and teeth (C) – for 8 test cases.

## 4. Discussion

This work presents a novel approach to the auto-segmentation of orodental structures that has the potential of supporting clinical decision-making in radiation therapy for patients with head and neck cancer. Advancing beyond existing orodental auto-segmentation methods^5–8^, our definition of mandible and maxilla sub-volumes is the first to account for variations in bone composition differences within these structures as well as laterality.

Establishing the boundaries between sub-volumes in the mandible and maxilla is challenging. The extent of the alveolar process varies across the mandible and maxilla structures^23^ and between patients^24^ and can be affected by dental extractions^14^. After dental extractions the size of the alveolar ridge decreases as it is resorbed and evolves into a so-called ‘residual ridge’ which is formed by denser cortical and trabecular bone with up to a 50% reduction in the height of the alveolar socket^14,25^. This process is typically more obvious in the molar and premolar areas, where teeth extractions are more common. For this study, a standardized alveolar process height of 3 to 5 mm was applied across all cases to define the alveolar sub-volumes of the maxilla and mandible. However, these dimensions represent a generalized approach and do not fully account for the variability inherent in individual anatomy and dental factors, which can significantly influence the structure and dimensions of the alveolar region, particularly in older edentulous patients. Future work will focus on refining our methodology to further advance towards a patient-specific approach to sub-volume definition and modeling.

Although the models achieved overall high overall segmentation performance, which directly translated into dosimetric accuracy, they exhibited limited applicability in segmenting teeth and sub-volumes often absent in the data (e.g., molars, premolars, maxilla alveolar sub-volumes). Missing data both in the training and test datasets may influence the performance of the models (supplemental Figure B2). Underrepresented structures in the training dataset clearly affected model performance, as the model cannot learn the features of the missing teeth or sub-volumes. On the other hand, underrepresentation of structures in the test dataset may affect reliability of the performance measures, as the computed metrics are based on limited data. A recent work by van Dijk et al. [ref RADMAP preprint here] has proposed a semi-automated teeth segmentation approach, the RADiation dose MAPping tool (RADMAP) tool, using an angular ray-based algorithm which is agnostic to missing teeth. While this tool is not fully automated and requires some manual adjustments on the angular rays defining the boundaries between teeth, it allows for domain knowledge input with regards to missing and shifted teeth. Future work will consider using the RADMAP approach to inform our DL teeth auto-segmentation model for a more generalized applicability.

Our mandible and maxilla sub-volumes definition aligns with the recently ASCO-endorsed ClinRad ORN staging system^16^, which incorporates radiological assessment of the vertical extent of bone damage, but also enables laterality determination. The clinical applicability of our models, however, expands to a wider range of clinical settings related but not exclusive to ORN staging. Automated sub-volume based assessments of pre-existing oral conditions (e.g., periodontal disease, caries) can support image-based early detection of bony changes in the mandible or maxilla resulting in more timely and granular interventions for reduced risk of more severe radiation-induced complications. On the other hand, a recent survey [reference Zeph’s physician review] has shown the poor performance of clinician-based radiographic diagnosis of mandibular damage leading to high variability in the application of the ClinRad ORN staging system, thus establishing a benchmark for more advanced computerized alternatives to the detection of pathological bony changes. Image-based features from computed tomography (CT) images have also been associated to ORN regions^26,27^. We have previously demonstrated that ORN^28^ or even earlier stages of mandible damage (e.g., microvascular damage)^29^ can be detected on magnetic resonance images (MRI); ongoing work is focused on black bone MRI for ORN detection^30^. Integrating our sub-volume auto-segmentation models into these image-based workflows for orodental damage detection will enhance the precision of the communication between the radiation oncology and dental teams.

In conclusion, this study presents a novel framework for DL-based auto-segmentation of orodental structures to develop a radiation-specific tool capable of spatially localizing dose-related differences in the jaw, thereby supporting a more effective approach to image-based bone damage detection, including ORN, and improving clinical decision-making in radiation oncology and dental care for head and neck cancer patients.

## Acknowledgements

This work was supported directly or in part by personnel funding/resource support from the National Institutes of Health (NIH) National Institute for Dental and Craniofacial Research (K01DE030524, U01DE032168, R21DE031082, R56/R01DE025248, R01DE028290); NIH National Cancer Institute (P01CA285249, K12CA088084, P30CA016672); the University of Texas MD Anderson Cancer Center Charles and Daneen Stiefel Center for Head and Neck Cancer Oropharyngeal Cancer Research Program; and the MD Anderson Image-guided Cancer Therapy Program. Additionally, M.A.N. received funding from the National Institutes of Health/National Institute of Dental and Craniofacial Research (NIH/NIDCR) through grant 1R03DE033550-01. C.D.F. and S.Y.L. received related funding support from the NIH/NIDCR (U01DE032168/ R01DE025248). C.D.F. also receives infrastructure and salary support through the NIH/NCI MD Anderson Cancer Center Core Support Grant (CCSG) Image-Driven Biologically-informed Therapy (IDBT) program (P30CA016672-47). S.Y.L. is supported through the CCSG Head and Neck Program (P30CA016672-48). K.A.W. was supported by an Image Guided Cancer Therapy (IGCT) T32 Training Program Fellowship from T32CA261856. L.V.V. received funding and salary support from KWF Dutch Cancer Society through a Young Investigator Grant (KWF-13529) and from NWO ZonMw through the VENI grant (NWO-09150162010173). K.K.B. and A.H.C. acknowledge support from the Image Guided Cancer Therapy Research Program at The University of Texas MD Anderson Cancer Center, which was partially funded by the National Institutes of Health/NCI under award number P30CA016672 and through a generous gift from the Apache Corporation. S.J.F. received funding from Hitachi, NIH/NCI, the National Association of Proton Therapy, Affirmed Pharma and NASA/Baylor College of Medicine and honoraria from IBA. N.W. was supported by a training fellowship from UTHealth Houston Center for Clinical and Translational Sciences T32 Program (Grant No. T32 TR004905) and a NIH National Institute of Dental and Craniofacial Research (NIDCR) Academic Industrial Partnership Grant (R01DE028290). S.J. acknowledges funding support from 1 R25 CA 265800-1 A1. Z.K. was supported by a doctoral fellowship from the Cancer Prevention Research Institute of Texas grant RP210042.

## Contributor Role Taxonomy (CRediT) Statement

Conceptualization: LHV, AHC, RH, CDF, MAN, ACM. Methodology: LHV, AHC, RH, LVV, KAW, AH, EW, RWA, CDF, MAN, ACM. Software: LHV, AHC, RH, CDF, MAN, ACM. Validation: LHV, AHC, RH, CDF, MAN, ACM. Formal analysis: LHV, AHC, RH, CDF, MAN, ACM. Investigation: LHV, AHC, RH, DJR, CW, HCW, CDF, MAN, ACM. Resources: DJR, HCW, MC, KKB, SYL, CDF, MAN, ACM. Data curation: LHV, AHC, RH, DJR, SJ, PG, NW, ZK, SM, JC, SA, AS, PO, AMSA, MAN, ACM. Writing - original draft: LHV, AHC, RH, CDF, MAN, ACM. Writing - review and editing: LHV, AHC, RH, LVV, DJR, CW, HCW, KAW, SJ, PG, NW, ZK, SM, JC, SA, AS, PO, AMSA, AH, EW, RWA, SJF, CEAB, KKB, MSC, MW, KAH, SYL, CDF, MAN, ACM. Visualization: LHV. Supervision: CDF, ACM. Project administration: ACM. Funding: CDF, KKB, MAN, ACM.

## Data Availability Statement

In accordance with the Final NIH Policy for Data Management and Sharing NOT-OD-21-013, data that support the findings of this study are openly available in an NIH-supported generalist scientific data repository (figshare) at https://doi.org/10.6084/m9.figshare.28615874 no later than the time of an associated publication; while public data is embargoed pending peer review, referees may access this data via private link at [link here] during the peer review process. Open access code deposition was placed in https://github.com/dr-mnaser/orodental-segmentation.

## Ethics Approval Statement

After institutional review board approval (RCR030800), data from a philanthropically funded observational cohort at The University of Texas MD Anderson Cancer Center (Stiefel Oropharynx Cancer Cohort, PA14-0947) were extracted for retrospective acquisition.

