## Supplementary Material for "Image-based Mandibular and Maxillary Parcellation and Annotation using Computer Tomography (IMPACT): A Deep Learning-based Clinical Tool for Orodental Dose Estimation and Osteoradionecrosis Assessment"

### Supplement A. Mandible and maxilla sub-volumes contouring

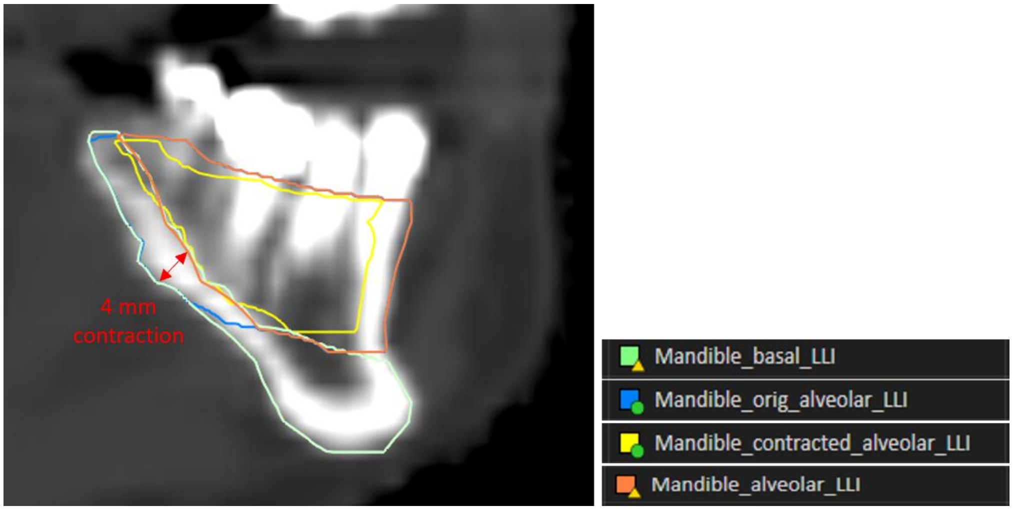


Figure A1. Example demonstrating a 4 mm contraction of the mandible contour at the molars level to allow for a differentiation between basal and alveolar bone regions.

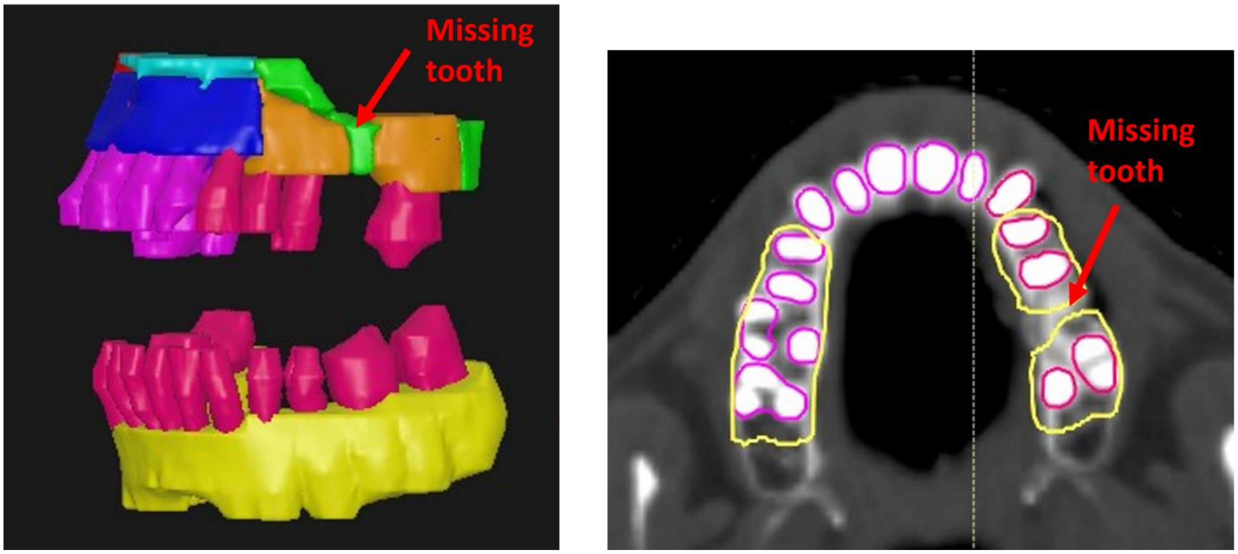


Figure A2. Example of a case with a missing tooth. To ensure generalizability of the model on edentulous or semi-edentulous patients, empty tooth sockets in cases with missing teeth were included in the alveolar region by manually adjusting the teeth expansion contour.

### Supplement B. Missing teeth analyses

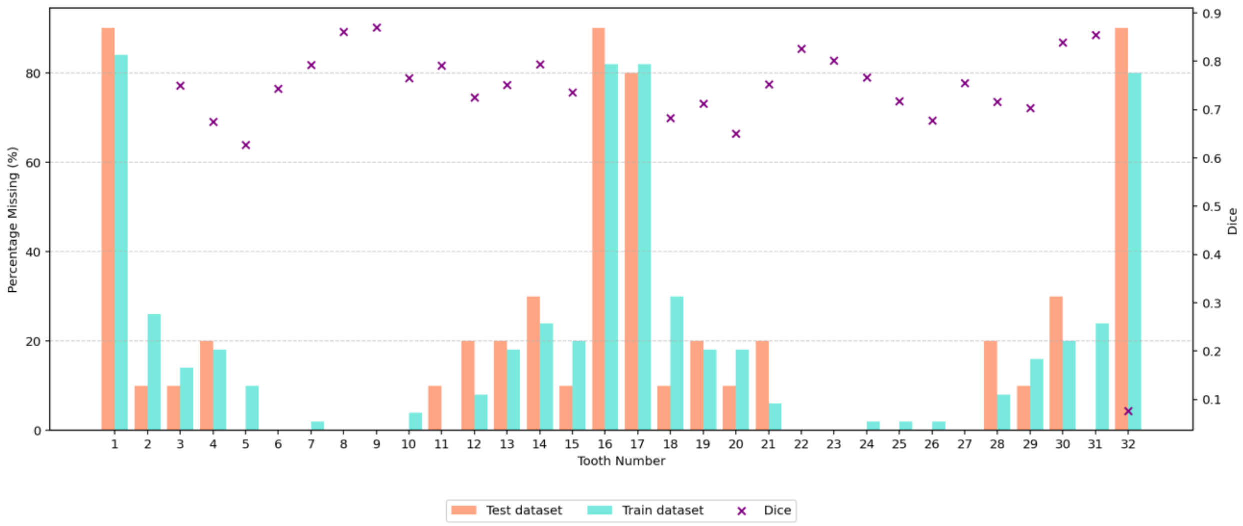


Figure B1. Bar chart illustrating the distribution of missing teeth percentages across the training and test datasets, with corresponding Dice metric values overlaid as purple markers. Teeth with higher missing percentages, such as the first and last molars (teeth 1, 16, 17, 32), exhibit lower Dice scores, indicating poorer segmentation performance in these regions. Conversely, teeth with lower missing percentages tend to have higher Dice scores, reflecting better segmentation accuracy.

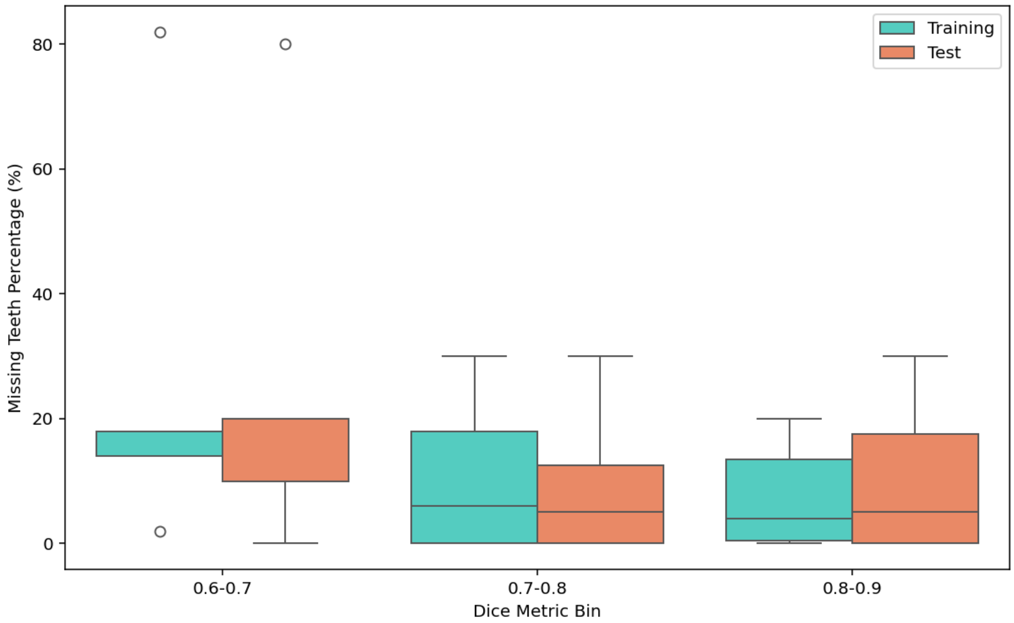


Figure B2. Boxplot showing the distribution of missing teeth percentages in training and test datasets, grouped by Dice metric bins. The results indicate an inverse relationship between Dice similarity and the percentage of missing teeth, with lower Dice scores (0.6–0.7) associated with higher missing teeth percentages. As Dice scores improve (≥0.7), the median and spread of missing teeth percentages decrease, suggesting better segmentation performance in cases with fewer missing teeth.

**Supplement C. Checklist for Artificial Intelligence in Medical Imaging (CLAIM)**

| Section / Topic | No. | Item | Page / Line | No | NA |
|---|---|---|---|---|---|
| TITLE / ABSTRACT | | | | | |
| | **1** | Identification as a study of AI methodology, specifying the category of technology used (e.g., deep learning) | **1** | | |
| ABSTRACT | | | | | |
| | **2** | Summary of study design, methods, results, and conclusions | **2** | | |
| INTRODUCTION | | | | | |
| | **3** | Scientific and/or clinical background, including the intended use and role of the AI approach | **3** | | |
| | **4** | Study aims, objectives, and hypotheses | **3** | | |
| METHODS | | | | | |
| *Study Design* | **5** | Prospective or retrospective study | **4** | | |
| | **6** | Study goal | **4** | | |
| *Data* | **7** | Data sources | **4** | | |
| | **8** | Inclusion and exclusion criteria | **4** | | |
| | **9** | Data pre-processing | **5** | | |
| | **10** | Selection of data subsets | **5/6** | | |
| | **11** | De-identification methods | | | |
| | **12** | How missing data were handled | **5/6** | | |
| | **13** | Image acquisition protocol | | | |
| *Reference Standard* | **14** | Definition of method(s) used to obtain reference standard | **4** | | |
| | **15** | Rationale for choosing the reference standard | **4** | | |
| | **16** | Source of reference standard annotations | **4** | | |
| | **17** | Annotation of test set | **4** | | |
| | **18** | Measures of inter- and intra-rater variability of features described by the annotators | | | |
| *Data Partitions* | **19** | How data were assigned to partitions | **6** | | |
| | **20** | Level at which partitions are disjoint | | | |
| *Testing Data* | **21** | Intended sample size | **6** | | |

| Section / Topic | No. | Item | Page / Line | No | NA |
|---|---|---|---|---|---|
| *Model* | **22** | Detailed description of model | **5/6** | | |
| | **23** | Software libraries, frameworks, and packages | **5/6** | | |
| | **24** | Initialization of model parameters | | | |
| *Training* | **25** | Details of training approach | **5/6** | | |
| | **26** | Method of selecting the final model | | | |
| | **27** | Ensembling techniques | | | |
| *Evaluation* | **28** | Metrics of model performance | **6** | | |
| | **29** | Statistical measures of significance and uncertainty | **6** | | |
| | **30** | Robustness or sensitivity analysis | | | |
| | **31** | Methods for explainability or interpretability | | | |
| | **32** | Evaluation on internal data | | | |
| | **33** | Testing on external data | **6** | | |
| | **34** | Clinical trial registration | | | |
| RESULTS | | | | | |
| *Data* | **35** | Numbers of patients or examinations included and excluded | **6/7** | | |
| | **36** | Demographic and clinical characteristics of cases in each partition | **6/7** | | |
| *Model performance* | **37** | Performance metrics and measures of statistical uncertainty | **7/8** | | |
| | **38** | Estimates of diagnostic performance and their precision | **10/11** | | |
| | **39** | Failure analysis of incorrect results | | | |
| DISCUSSION | | | | | |
| | **40** | Study limitations | **11/12** | | |
| | **41** | Implications for practice, including intended use and/or clinical role | **11/12** | | |
| OTHER INFORMATION | | | | | |
| | **42** | Provide a reference to the full study protocol or to additional technical details | | | |
| | **43** | Statement about the availability of software, trained model, and/or data | **1** | | |
| | **44** | Sources of funding and other support; role of funders | | | |
